# Projected sudden step-wise increase in risk of large measles outbreaks in the Netherlands as susceptible children enter secondary school in 2025/26

**DOI:** 10.1101/2025.08.27.25334424

**Authors:** James D Munday

## Abstract

**Background:** Many unvaccinated children were naturally immunised through infection during the last national measles epidemic in the Netherlands (2013/14). Over time susceptibility amongst school aged children in key under-vaccinated populations is likely to increase and warrants investigation.

**Methods:** We constructed a network to reflect the risk of transmission within and between schools in the Netherlands. Estimating school-level susceptibility rates using historical estimates and vaccine uptake. We evaluated the annual outbreak risk since 2014 projecting to 2030.

**Results:** As children born since the 2013/14 outbreak enter secondary school, the connectedness of the school-mediated contact network between susceptible children will increase sharply, resulting in a sudden increase in the risk of large measles outbreaks.

**Conclusions:** We anticipate a substantial increase in the risk of large measles outbreaks mediated by school-based transmission in 2025 and 2026 as larger numbers of susceptible children enter secondary school.

## Introduction

The Netherlands has historically experienced large measles epidemics, despite high uptake of the highly effective Measles, Mumps and Rubella (MMR) vaccine (>95%). Notably, since the introduction of the measles vaccine in 1976, there have been large outbreaks of measles with intervals of between 10 and 13 years (1976/7, 1988 [1], 1999/2000 [2] and 2013/4 [3]). The two largest outbreaks since the introduction of MMR in 1987 (>2700 reported cases with estimated total infection count of ca. 30,000) were both strongly associated with the orthodox Protestant population, which have collective MMR uptake of approximately 25% [4]. Estimates suggest that the epidemic in 2013/14 was sufficiently large to effectively immunise all susceptible children in the orthodox Protestant community. Over time, however, the immunity in this population is expected to have declined as new children are born and remain unvaccinated as they become more socially engaged. This in turn would be expected to increase the risk of large epidemics again. In addition, the latest national estimates indicate falling MMR uptake in other key socio-religious groups [5], correlating with a notable increase in measles case reports (including 202 cases in 2024) [7]. Recent outbreaks affected unvaccinated children through transmission within families, care and educational settings, however, no cases were reported in the orthodox Protestant community. These combined factors place anticipation of measles outbreaks high on the public health agenda.

Recent work shows that the geographical signature and size of previous epidemics is well explained by school-based outbreaks linked by transmission between children attending separate schools and living in shared households [6]. It was demonstrated by analysing a ‘network’ of schools, that orthodox protestant schools form strongly connected clusters, generating concentrated pockets of unvaccinated children. In this manuscript we examine the evolution of epidemic risk using a similar school-household network approach, with a focus on the orthodox Protestant community.

### Methods

Following the approach of Munday et al. (2024), we constructed a network of schools using data from the Dutch National Education Executive (DUO) by counting the number of contact pairs between each pair of schools (Supplementary Material and Munday et al. 2024 [6]). To assess the risk of outbreaks on the network in each year following the 2013 outbreak, we estimated the immunity profile of each school by replenishing the susceptible population in schools as they mature to each school year (Figure 1, Supplementary Material). Concretely, we assumed that the 2013/14 outbreak resulted in 100% immunity in schools in 2014. For simplicity we assume that all children born prior to September 2014 also have acquired measles immunity, either through vaccination or by natural infection. Hence, for 3 years following the outbreak we expect no susceptible children to begin school. In the 4th year we assume children entered the first year of each primary school with the same MMR uptake as estimated in 2013. For subsequent years we continued to replenish susceptible children in each school year such that by the 11th year primary schools were returned to their 2013 level of immunity profile. In the 12th year children entered the first year of each secondary school with the same immunity profile as estimated for 2013. We continued such that by the 17th year, primary and secondary schools were returned to their 2013 immunity profile. For each of the annual scenarios we constructed a network of transmission potential, where vertices represent schools (for example schools i and j) and the edges between them quantify the probability of an outbreak in school i causes an outbreak in school j through transmission between students at home.

**Figure 1.**
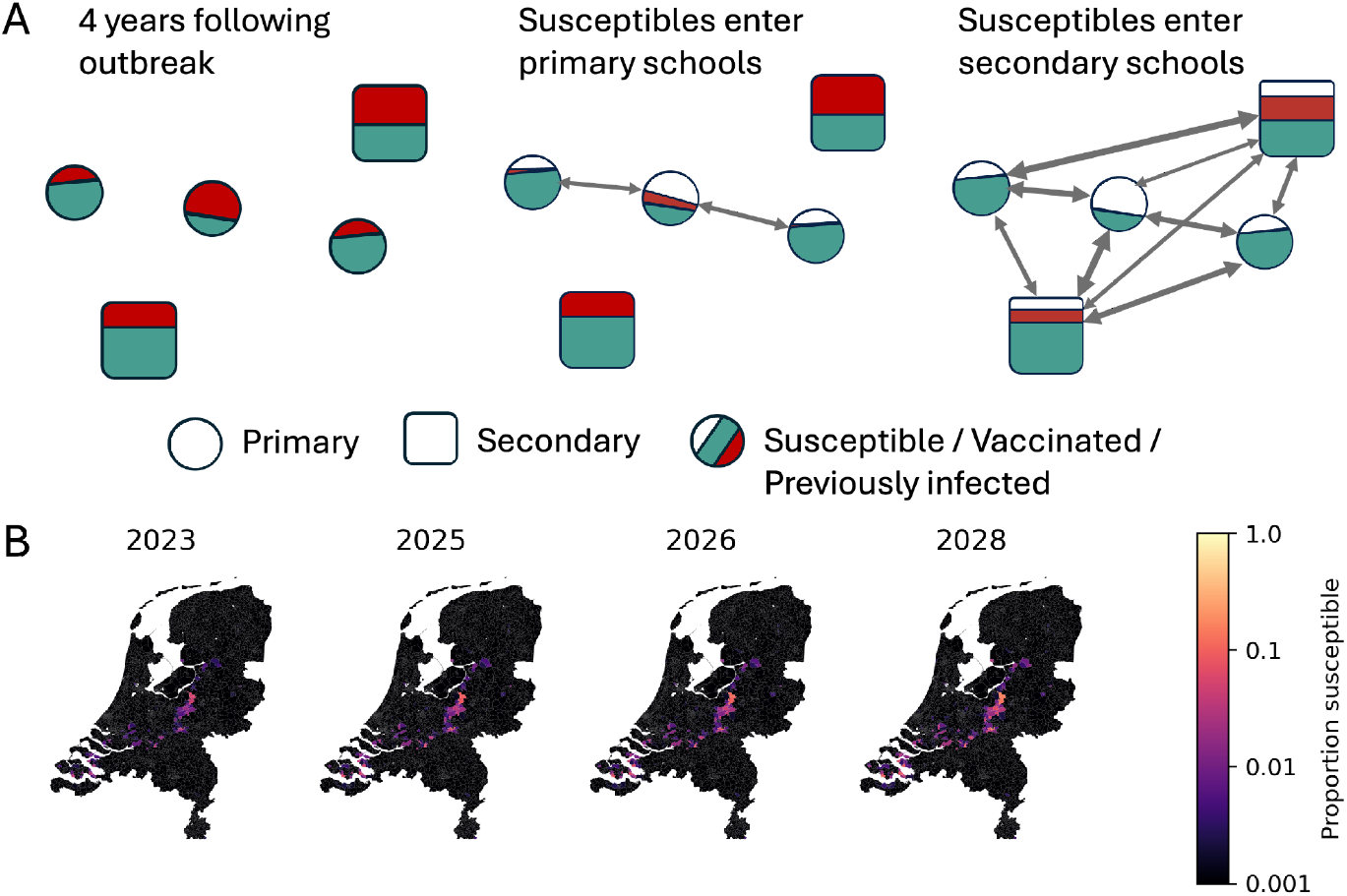
The progression of susceptible children through the school system after a large outbreak that provides near complete natural immunity to unvaccinated children. A) model of replenishment of susceptible children in schools over time. Circles represent primary schools, and squares secondary schools. Green represents the proportion of the school vaccinated, red represents the proportion of the school with naturally acquired immunity, white represents the proportion of the school susceptible to measles infection. The arrows represent possible transmission paths between schools. B) Choropleths showing geographic distribution of susceptibility in schools in the phase where susceptible children enter secondary school.

To evaluate the connectedness of the network as susceptible children progress through the school system, we first calculated the spectral radius of the network’s adjacency matrix, which provides a measure of overall connectedness of susceptible children. Secondly, we simulated outbreaks seeded from each orthodox-protestant school, and propagated the outbreak along the transmission probability network as presented in Munday et al. (2024) (summary in Supplementary Material). From the outbreaks we quantified the number of schools affected and the number of children infected, under the assumption that school outbreaks reach a theoretical final size [8].

## Results

The spectral radius increases steadily for the first eight years after susceptible children enter school, suggesting susceptible children gradually become more connected on the network (Figure 2 A). The increase accelerates when susceptible children enter secondary schools, suggesting that susceptible children become much more connected.

**Figure 2.**
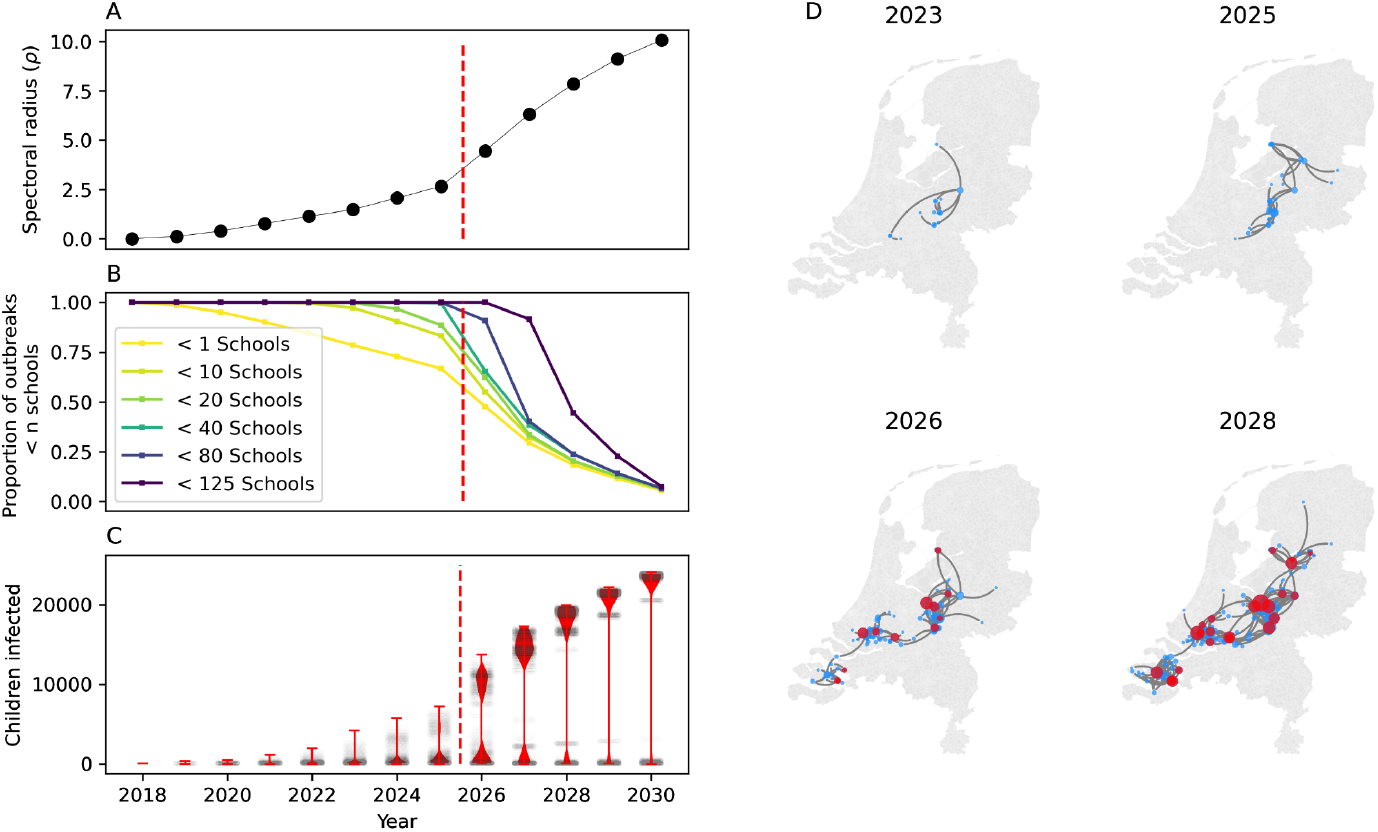
The evolution of measles outbreak risk on the school-household network in years since 2018. A) The spectral radius of the transmission probability network for each year. B) The proportion of outbreaks seeded in orthodox protestant schools that exceed final size thresholds of 1 to 125 schools. C) The distribution of the number of children infected in outbreaks simulated on the network, seeded in orthodox protestant schools. D) Graph visualisations showing examples of largest out-components (outbreaks) for individual edge percolation instances for years 2023 to 2028, blue vertices show primary schools, red vertices show secondary schools (geographically accurate).

We found that the number of schools affected by each outbreak increased year on year (Figure 2 B). However, larger outbreaks of greater than 80 schools, in particular, jumped from having a relatively low risk in the year that susceptible children enter primary school (i.e. maximum susceptibility in all primary schools), to over 50% risk in the year that susceptible children first entered secondary schools.

We estimate that this jump in the number of schools affected increases the expected overall epidemic size from several hundred infections clustered in small geographic areas, to several thousand infections spread widely across the country (Figure 2 C and D). In the years that follow, the expected number of infections steadily increases, as susceptible children accumulate in connected schools (Figure 2 C).

All three of these observations indicate that the network becomes more connected as children enter secondary school, highlighting their key role in forming bridges between pockets of susceptible children within primary schools, ultimately facilitating large outbreaks. This change is also reflected in the offspring distribution of primary and secondary schools (Supplementary figure S4), where outbreaks in primary schools rarely seed new outbreaks in more than 3 or 4 adjacent schools, but secondary schools can seed new outbreaks in as many as 25 adjacent schools over the course of an outbreak.

## Discussion

Our findings suggest that the Netherlands is approaching a period of elevated risk of large measles outbreaks as children born since the last major measles epidemic (ending in 2014) enter secondary school in the next two years. The sharp rise in outbreak risk in September 2026 is due partly to our assumption that all children born before 2014 are immune. This is a simplification and this effect is likely to be staggered over a number of years with the greatest effects in September 2025 and September 2026. Nevertheless, the substantial rise in risk we demonstrate highlights the need to monitor vaccine-uptake and susceptibility profiles on the basis of social networks as well as their geographical distribution.

The framework could be improved with the use of up-to-date vaccine registry and school records data, where we currently induce vaccine coverage by school and school network structure from 2013 data. RIVM is working with the Dutch Education Ministry (DUO) and the Central Statistics office (CBS), undertaking the necessary steps to overcome these limitations.

We advocate for governmental administrative data to be used to facilitate surveillance and response efforts, providing a way to inform of transmission risk and identify areas for targeted surveillance and intervention efforts. We emphasise the need to understand how the changing vaccine uptake patterns in community sub-groups, and promote use of empirical network analysis to anticipate future challenges with measles outbreaks in the Netherlands and other settings with declining MMR uptake.

## Data Availability

The code and data used in this study are available on github

https://github.com/jdmunday/InterepidemicPeriodsNL

## Data Availability

The code and data used in this study are available on github

https://github.com/jdmunday/InterepidemicPeriodsNL

## Data Availability

The code and data used in this study are available on github

https://github.com/jdmunday/InterepidemicPeriodsNL

## Data Availability

The code and data used in this study are available on github

https://github.com/jdmunday/InterepidemicPeriodsNL

## Data Availability

The code and data used in this study are available on github

https://github.com/jdmunday/InterepidemicPeriodsNL

## Ethical statement

The analysis in this manuscript uses only open access data.

## Data availability

The code and data used in this study are available on github: https://github.com/jdmunday/InterepidemicPeriodsNL

## Acknowledgements

I am grateful to the Dutch National Institute for Public Health and the Environment (RIVM) for their fruitful collaboration especially to Susan Hahné, Tom Woudenberg, Jacco Wallinga, Don Klinkenberg and Albert Jan van Hoek who each provided detailed support. I also wish to thank Katherine Atkins for her previous support in developing the approach used in the model. Finally, I would like to thank the Office of Education (DUO) for their support with data access, particularly Erik Fleur and Marc Meurs.

