## Supplementary Materials for "Projected sudden step-wise increase in risk of large measles outbreaks in the Netherlands as susceptible children enter secondary school in 2025/26"

### Additional Methods

#### Constructing the school-household network

*For full details, please refer to Munday et al. (2024) [1]. Here we provide a short summary of the approach.*

We constructed a network of schools using data from the Dutch National Education Executive (DUO) by counting the number of contact pairs between each pair of schools in each household represented in the data. We constructed a network of schools as linked through household contacts. Where an edge between school i and j is the number of unique child to child contacts within a set of households, $k$,

$$C_{i,j} = \sum_k (n_{k,i} n_{k,j}) \qquad (1)$$

where $n_{k,i}$ is the number of children in household k that attend school i. We estimated the probability, $P_{trans,i,j}$ of a measles outbreak in school *i* spilling over to school *j* based on the school network, the reproduction number of measles ($R_0$) and the school level vaccine uptake in schools, estimated by Klinkenberg et al. (2022) [2]in the same way as Munday et. al 2024[1].

$$P_{trans,ij} = 1 - (1 - P_j^I P_i^S q P_i^{OB})^{C_{ij}} \qquad (2)$$

Where, $P_j^I$ and $P_i^{OB}$ are found by solving for the theoretical final outbreak size $R(\infty)$ in school j and i respectively for a school level immunity coverage $V_j$.

$$R_j(\infty) = (1 - V_j)(1 - e^{-(1-V_j)R_0 R(\infty)}) \qquad (3)$$

$P_i^S$ is equal to the estimated proportion susceptible and $q$ is the probability of transmission between an infected and susceptible child who live in the same household.

#### Propagating susceptible children through the school system

We estimated the school-level immunity profile in each year following the major outbreak of 2013-2014 by altering the school-specific susceptible proportion based on the number of students who we estimated to be immune either through vaccination or via prior infection. We assume that all unvaccinated children were effectively immunised during the 2013-2014 outbreak. We assume that as children are born they are vaccinated at the same rates as prior to the 2013 outbreak. For the following 4 years we assume that no susceptible children enter the school system. At year 4 we replenish 1/8th of the susceptibles that were estimated to be in each primary school in 2013, according to Klinkenberg et al [2]. In year 5 we add another 1/8th. We continue until year 12, when the immune profile of primary schools is the same as before the outbreak in 2013 - and immunity is assumed to be acquired only by vaccination. In year 13 we replenish 1/5 of the susceptibles that were estimated to be in each primary school in 2013 and repeat each year until the secondary schools have the same profile as estimated prior to the epidemic in 2013.

**Percolation probability based outbreak simulations**

To estimate the extent of outbreaks on the network we sampled network edges with the probability assigned by the transmission probability network described by equation 2. We identified ‘outbreaks’ on the resulting networks as successive chains of out edges (out-components) of each school in the network (Figure S1).

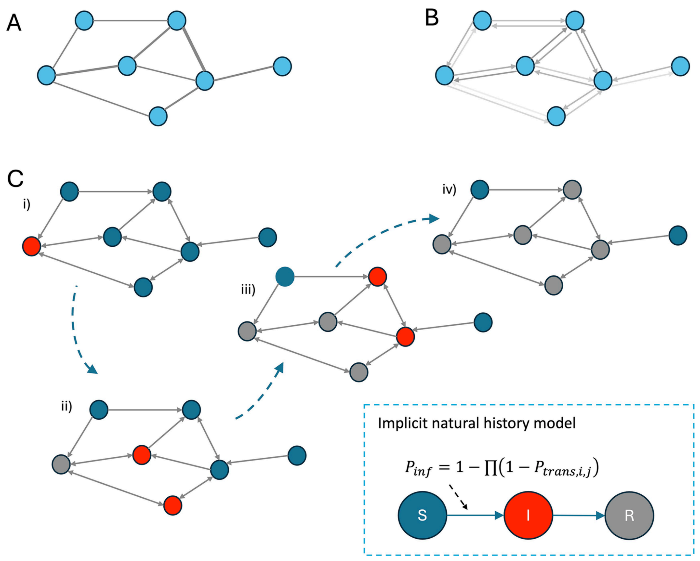


**Figure S1:** (A) Shows the contact network between schools (blue circles) where edge weights, indicated by thickness, give the number of unique contact pairs though shared households. (B) Shows the transmission probability network where the directed edge weights give the probability of transmission between schools in each direction, as calculated in Eq 2. (C) Shows binary outbreak networks where directed edges are given a value of 1 or 0, successive generations of an outbreak are shown in panels (i) to (iv), where schools occupy one of 3 epidemiological states (susceptible (S), infected (I), and recovered (R)). In each generation schools connected by an out edge (weight 1) from an infected school are infected in the next generation of the model. This process continues until no outages from infected schools reach susceptible schools. This figure and caption are lifted and unedited from[1]

The ‘largest component’ was determined as the out-component with the highest number of schools.

We estimated the number of children infected in each ‘outbreak’ by taking the deterministic final size estimate per school as described in equation 3 and summing the number of students affected over all schools in the component.

### Additional figures

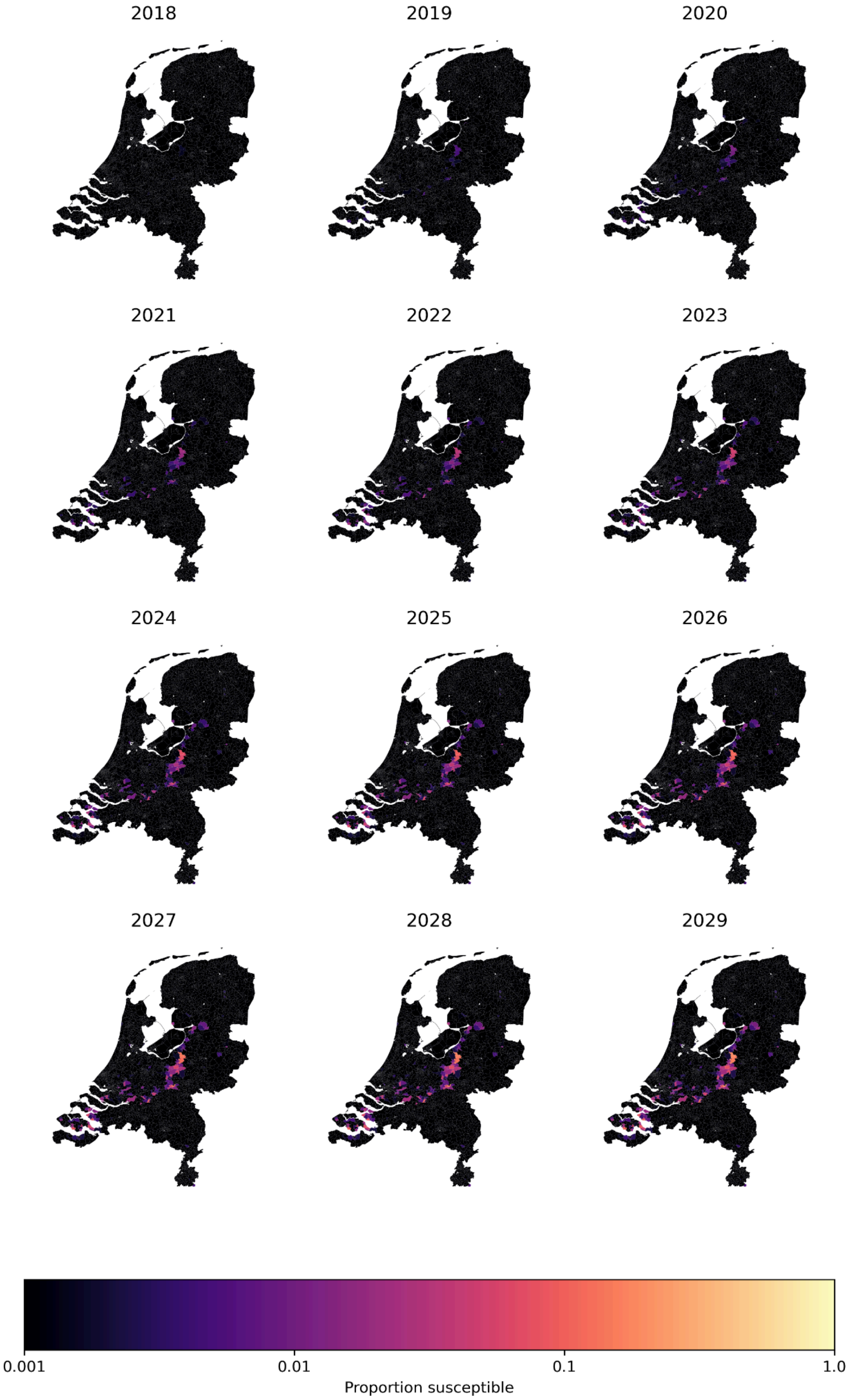


**Figure S2:** the proportion of school children resident in each post-code 4 area who are modelled as susceptible to measles in each year after the first susceptibles enter primary school (4 years after previous outbreak).

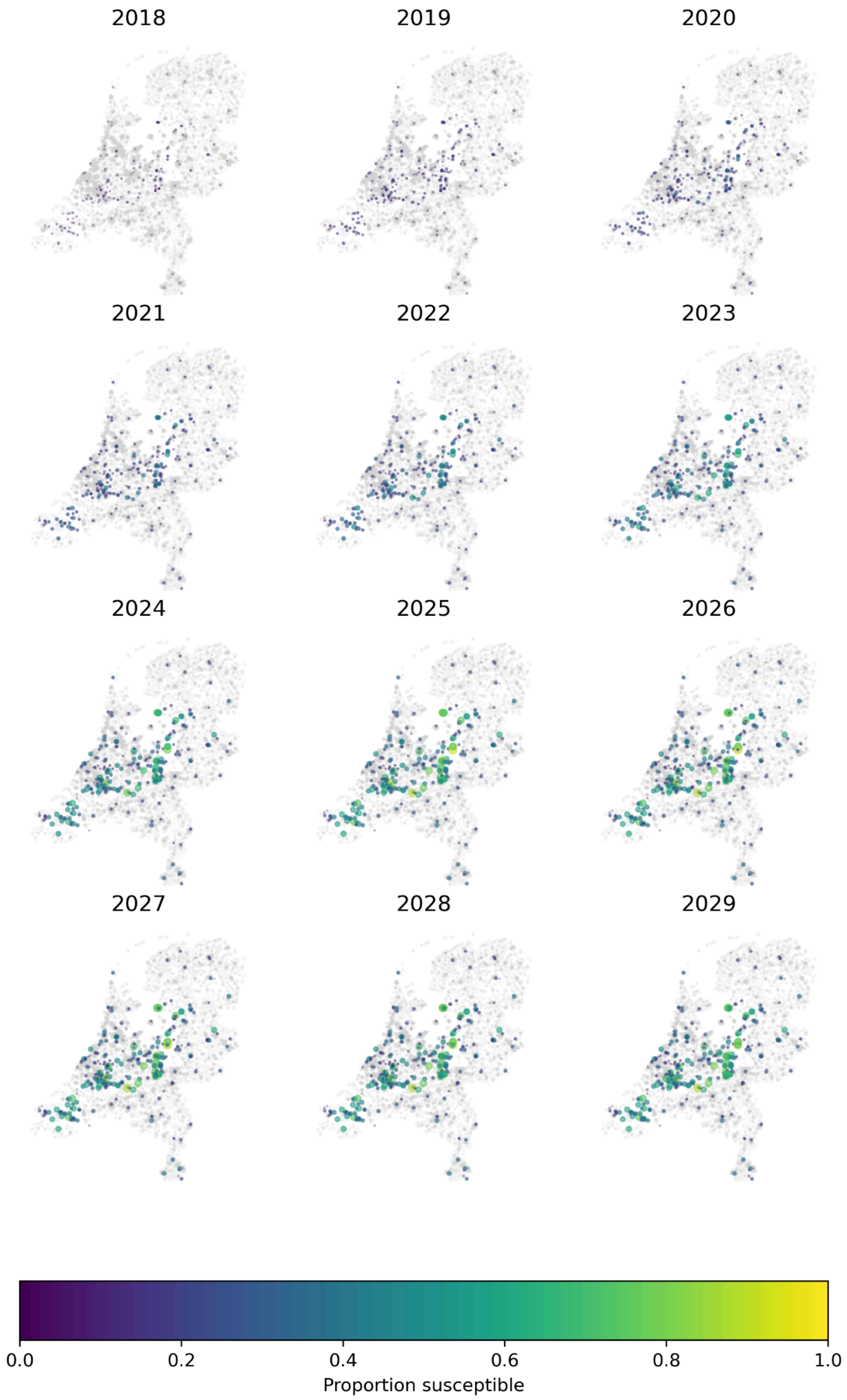


**Figure S3:** The proportion of school children attending each school who are modelled as susceptible to measles in each year after the first susceptibles enter primary school (4 years after previous outbreak). Points show the location of schools, hue and size represent the proportion of the school that is susceptible to measles.

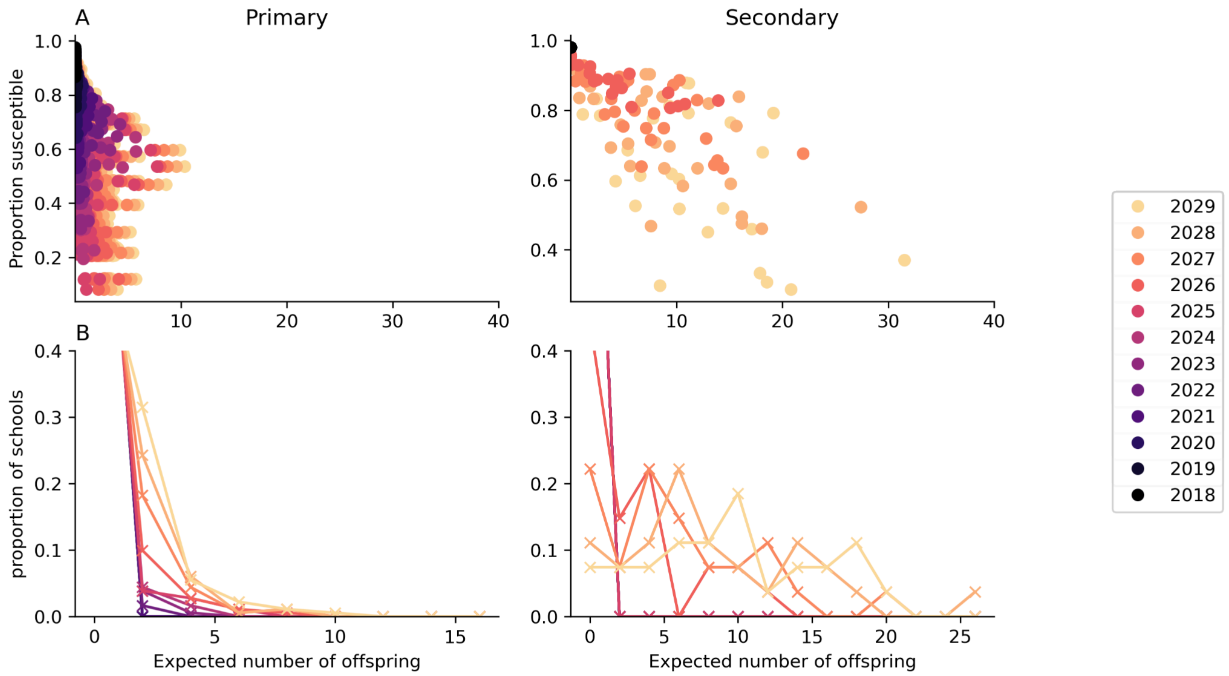


**Figure S3:** The average number of offspring from each school if infected (the weighted degree of the transmission probability network) divided into primary and secondary school components. A) Scatter plot of the expected offspring by the proportion of the school that is immune. B) Offspring distributions over the entire network.
